# Screening for Probable Undiagnosed Hypertension in US Adults Using Interpretable Machine Learning: An NHANES 2017-2018 Study

**DOI:** 10.64898/2026.06.23.26356178

**Authors:** Mukund Thakur, Sanjaya Aacharya, Abhigan Babu Shrestha, Pranjal Tiwari, Roshan Sah, Pratik Yadav, Dhrub Yadav, Binay Thakur

## Abstract

**Background:** Hypertension remains one of the most challenging healthcare problems in the community. It is a common, measurable, and treatable condition that is nonetheless responsible for millions of preventable deaths each year. Hypertension affects approximately 1.28 billion adults worldwide, yet fewer than half (46%) are aware of their condition. Undiagnosed hypertension is a critical public health gap, contributing to preventable cardiovascular morbidity through silent end-organ damage. Machine learning can enable community-level screening using routinely available, non-invasive data, without the requirement of laboratory investigations.

**Methods:** We conducted a cross-sectional ML study using the National Health and Nutrition Examination Survey (NHANES) 2017-2018 cycle. Adults aged ≥18 years were included; those with a self-reported prior hypertension diagnosis (n=1,930) were retained as negative controls. Probable undiagnosed hypertension was defined as mean blood pressure ≥130/80 mmHg among participants reporting no prior hypertension diagnosis, consistent with the ACC/AHA 2017 threshold. Three classifiers: Logistic Regression (LR), Random Forest (RF), and Extreme Gradient Boosting (XGBoost) were trained on eight non-invasive predictor variables. Performance was assessed using AUC-ROC, sensitivity, specificity, and F1 score with bootstrap 95% confidence intervals (CIs). Stratified 5-fold cross-validation was applied.

**Results:** Of 9,254 NHANES 2017-2018 participants, 5,237 adults were included after exclusion criteria were applied; 1,072 (20.5%) had probable undiagnosed hypertension. LR achieved the highest test AUC of 0.611 (95% CI: 0.571-0.652), with sensitivity 0.535 (95% CI: 0.473-0.600) and specificity 0.594 (95% CI: 0.560-0.626). RF demonstrated near-absent sensitivity (0.047) despite adequate AUC (0.607), while XGBoost performed intermediately (AUC: 0.599; sensitivity: 0.279). Diabetes status, sex, and age were the most influential predictors by permutation feature importance.

**Conclusion:** ML classifiers trained on eight non-invasive variables demonstrated modest but consistent discrimination for identifying probable undiagnosed hypertension, supporting the feasibility of laboratory-free community screening. External validation in diverse populations is warranted before clinical implementation.

## INTRODUCTION

Despite being readily detectable and effectively treatable, hypertension continues to account for a substantial share of preventable cardiovascular mortality worldwide. Approximately 1.28 billion adults worldwide live with hypertension, yet fewer than half (46%) are aware of their condition.^1^ Hypertension is one of the major modifiable risk factors for stroke, myocardial infarction, heart failure, and chronic kidney disease. Although effective and inexpensive treatments exist, uncontrolled hypertension continues to drive a substantial share of cardiovascular mortality, a burden that falls disproportionately on low- and middle-income countries with limited diagnostic infrastructure.^2^

Undiagnosed hypertension is a major, often overlooked challenge within the broader epidemic called ‘Hypertension’. By lowering the diagnostic threshold to ≥130/80 mmHg, the 2017 ACC/AHA guidelines captured more people as hypertensive, which in turn increased the number of individuals who remain unaware of their condition.^3^ Because it often progresses without symptoms, untreated hypertension can lead to severe end-organ damage, such as left ventricular hypertrophy, nephrosclerosis, and hypertensive retinopathy. Because conventional screening depends on a clinical encounter, it remains inaccessible to populations with limited healthcare access.

Machine learning (ML) has become a powerful tool for identifying disease risk from routinely collected health data. Machine learning has been applied to hypertension risk prediction in several NHANES-based studies,^4,5^ although a systematic review of this literature indicates that most published models depend on laboratory or clinical variables unavailable in community settings.^6^ Most published models require data from laboratory investigations like lipid panels, fasting blood glucose, or renal function markers to screen for hypertension. This limits their deployment in primary care or community settings without laboratory access. Moreover, most prior studies target general hypertension risk prediction rather than the specific, clinically urgent question of identifying adults who are already hypertensive but are unaware of their diagnosis.

No prior study has specifically targeted probable undiagnosed hypertension — defined as objective blood pressure elevation accompanied by self-reported unawareness — using the 2017 ACC/AHA threshold and a predictor set restricted to non-invasive, laboratory-free variables in the NHANES 2017–2018 cycle. An interpretable screening model based on routinely observable variables would extend cardiovascular risk identification to settings where laboratory resources are limited.

Therefore, this study aimed to: (1) develop and compare interpretable ML classifiers for screening probable undiagnosed hypertension among US adults using NHANES 2017–2018 data; (2) restrict predictor variables to eight routinely collectable, non-invasive features; and (3) identify the most influential predictors of undiagnosed hypertension status using permutation feature importance analysis.

## METHODS

### Study Design and Data Source

We carried out a cross-sectional ML study using publicly available secondary data from the NHANES 2017–2018 cycle (J cycle). NHANES is a nationally representative, ongoing survey conducted by the Centers for Disease Control and Prevention (CDC) to assess the health and nutritional status of non-institutionalised US adults through standardised interviews, physical examinations, and laboratory tests.^7^ All component data files are freely accessible at: https://wwwn.cdc.gov/nchs/nhanes/. This study used entirely anonymised, publicly available data and required no additional institutional ethics approval.

### Study Population

#### Inclusion criteria

We included adults aged 18 or older who had both recorded blood pressure measurements and a clear answer to the hypertension awareness question (BPQ020).

#### Exclusion criteria

We excluded participants with age <18 years, invalid or missing blood pressure awareness response (BPQ020 not coded 1 or 2), and missing blood pressure measurements. Of 9,254 NHANES 2017–2018 participants, 3,398 were excluded for age <18 years, 10 for invalid blood pressure awareness responses, and 609 for missing blood pressure measurements, yielding a final sample size of 5,237 adults (Figure 3). Of this sample, 1,930 participants with a known hypertension diagnosis were retained as negative controls, as their status precluded classification as undiagnosed.

### Outcome Definition

The primary outcome was *probable undiagnosed hypertension*, defined as:

1. Mean systolic blood pressure ≥130 mmHg OR mean diastolic blood pressure ≥80 mmHg, calculated from up to three standardised oscillometric readings (BPXSY1–3, BPXDI1–3), consistent with the ACC/AHA 2017 hypertension threshold;^3^ AND
2. Participant self-report of never having been informed by a healthcare professional of a hypertension diagnosis (BPQ020=2).

Participants satisfying both criteria were classified as positive (outcome=1). All others — including previously diagnosed hypertensives (BPQ020=1) who served as negative controls — were classified as negative (outcome=0). In the NHANES protocol, a reading of 0 mmHg indicates a measurement failure rather than a real physiological value. To avoid artificially lowering our blood pressure estimates, we replaced these ‘0’ readings with missing values before calculating the mean.

### Predictor Variables

Eight non-invasive predictor variables were selected based on clinical relevance, prior literature, and suitability for community-based screening without laboratory access:

1. age (RIDAGEYR, years),
2. sex (RIAGENDR),
3. body mass index (BMXBMI, kg/m²),
4. waist circumference (BMXWAIST, cm),
5. smoking status (SMQ020: ever smoked ≥100 cigarettes in lifetime),
6. diabetes status (DIQ010: doctor-confirmed diabetes diagnosis),
7. vigorous physical activity (PAQ605: vigorous work activity in past week),
8. weekday sleep duration (SLD012, hours).

Sleep duration was included given its established association with nocturnal blood pressure changes and cardiovascular risk. All variables are self-reported or anthropometrically measured and require no laboratory investigation.

### Data Processing

The eight NHANES component files were merged on the unique participant sequence number (SEQN). To handle missing data, we applied median imputation for continuous variables and mode imputation for categorical ones. Categorical variables were binary-encoded (1=positive category, 0=otherwise) before modelling. Because negative cases significantly outnumbered the positive ones, we addressed this class imbalance using class-weighted training. This ensures the models prioritize identifying the minority positive cases, using ‘class_weight=“balanced”’ for Logistic Regression and Random Forest, and adjusting ‘scale_pos_weight’ for XGBoost.

### Machine Learning Approach

The dataset was partitioned into training (80%) and test (20%) sets using stratified random sampling to preserve class distribution (random_state=42). Feature standardisation using StandardScaler was fitted exclusively on training data and applied to the test set to prevent data leakage. Stratified 5-fold cross-validation was applied to the training set to estimate generalisation performance. This gives us a more reliable estimate of how the model will perform on new, unseen data. After the cross-validation, the models were trained on the full 80% of the training dataset, and later evaluated on the remaining 20% of the testing dataset.

Three classifiers were evaluated:

1. **Logistic Regression (LR):** LR is a standard baseline model valued for its transparency in explaining how specific variables influence outcomes;
2. **Random Forest (RF):** RF makes predictions by aggregating the decisions of 100 individual decision trees trained on bootstraped samples; and
3. **Extreme Gradient Boosting (XGBoost):** XGBoost is a gradient boosting framework that builds 100 decision trees sequentially, each correcting residual errors from the previous one.

All models used default hyperparameter configurations except for class imbalance adjustments described above.

Default configurations were deliberately retained to ensure that observed performance differences across different ML models reflect each algorithm’s inherent characteristics rather than the effects of extensive tuning. This choice also reduces the risk of overfitting to this specific training sample.

### Model Evaluation

The primary metric was AUC-ROC. Secondary metrics were sensitivity (recall for the positive class), specificity (recall for the negative class), and F1 score. All metrics are reported as point estimates with 95% bootstrap confidence intervals derived from 1,000 stratified resamples with replacement. Resamples in which only one class was represented were excluded, as the AUC could not be computed in their absence.

LR was selected as the optimal model based on its highest test AUC (0.611) combined with superior sensitivity (0.535) and F1 score (0.344), making it the most clinically balanced classifier for a screening application where sensitivity is the primary objective.

### Interpretability Analysis

Permutation feature importance was applied to the best-performing model (LR), quantifying the mean decrease in test set AUC when each feature was randomly permuted across 30 independent repeats. Results are expressed as mean ± standard deviation. This technique ranks predictors by their actual contribution to discrimination, rather than by statistical significance alone.

### Software and Reporting

All analyses were performed in Python 3.13 using scikit-learn,^8^ XGBoost, pandas, NumPy, and Matplotlib. The complete analysis pipeline is available from the corresponding author upon reasonable request. This study adhered to the Transparent Reporting of a Multivariable Prediction Model for Individual Prognosis or Diagnosis (TRIPOD) reporting guidelines.^9^ As NHANES employs a complex multistage probability sampling design, sampling weights were not applied to the ML pipeline; population-level prevalence estimates should therefore be interpreted with caution. Consequently, our findings are representative of the study cohort (NHANES dataset) rather than the weighted U.S. population, which may affect the generalizability of our results.

## RESULTS

### Study Population

Of 9,254 NHANES 2017–2018 participants loaded across eight component files, 5,237 adults met the inclusion and exclusion criteria and comprised the final analytic sample (Figure 3). A total of 1,072 participants (20.5%) were classified as having probable undiagnosed hypertension (positive class), and 4,165 (79.5%) served as the negative comparison group — comprising 1,930 previously diagnosed hypertensives retained as negative controls and 2,235 participants with blood pressure below the ACC/AHA threshold. Participant characteristics stratified by outcome status are presented in **Table 1**.

**Table 1.** Baseline characteristics of included participants stratified by probable undiagnosed hypertension status (NHANES 2017–2018). Continuous variables: mean ± SD; categorical variables: n (%). p-values from Welch’s independent samples t-test (continuous) and Pearson’s chi-square test (categorical). Two-sided tests applied throughout.

| Variable | Overall<br>(N=5,237) | Probable<br>Undiagnosed<br>HTN (n=1,072) | No<br>Undiagnosed<br>HTN (n=4,165) | p-value |
| --- | --- | --- | --- | --- |
| Age (years) | 49.9 ± 18.6 | 52.7 ± 16.6 | 49.1 ± 19.0 | <0.001 |
| BMI (kg/m<sup>2</sup>) | 29.7 ± 7.3 | 30.0 ± 7.9 | 29.6 ± 7.2 | 0.099 |
| Waist Circumference (cm) | 100.1 ± 16.8 | 101.3 ± 17.0 | 99.8 ± 16.7 | 0.009 |
| Sleep Duration (hours) | 7.6 ± 1.7 | 7.5 ± 1.7 | 7.6 ± 1.6 | 0.133 |
| Female Sex, n (%) | 2,696 (51.5%) | 466 (43.5%) | 2,230 (53.5%) | <0.001 |
| Ever Smoked ≥100 Cigarettes, n (%) | 2,133 (40.7%) | 436 (40.7%) | 1,697 (40.7%) | 0.993 |
| Diabetes Status (Yes), n (%) | 793 (15.1%) | 108 (10.1%) | 685 (16.4%) | <0.001 |
| Vigorous Physical Activity, n (%) | 1,274 (24.3%) | 278 (25.9%) | 996 (23.9%) | 0.182 |

Participants with probable undiagnosed hypertension were significantly older than those in the negative class (mean age 52.7 ± 16.6 vs. 49.1 ± 19.0 years; p<0.001). Among anthropometric measures, waist circumference differed significantly between groups (101.3 ± 17.0 vs. 99.8 ± 16.7 cm; p=0.009), whereas BMI did not reach statistical significance (30.0 ± 7.9 vs. 29.6 ± 7.2 kg/m²; p=0.099), suggesting that central adiposity may be a more sensitive marker of undiagnosed hypertension than overall adiposity. Female sex was significantly less prevalent in the positive class (43.5% vs. 53.5%; p<0.001), indicating a higher burden of undiagnosed hypertension among male participants. Diabetes status was significantly less prevalent in the positive class (10.1% vs. 16.4%; p<0.001), reflecting greater healthcare utilisation among diabetic individuals and the consequent likelihood of prior hypertension recognition. Smoking prevalence was identical across both groups (40.7%; p=0.993). Sleep duration and vigorous physical activity did not differ significantly between groups (p=0.133 and p=0.182, respectively).

### Cross-Validation Performance

We assessed our three models (Logistic Regression, Random Forest, and XGBoost) using five-fold stratified cross-validation. All models yielded similar results, with AUC estimates of 0.616 ± 0.019 (LR), 0.612 ± 0.006 (RF), and 0.621 ± 0.013 (XGBoost) (Table 2), and their low standard deviations indicate good stability without general overfitting. A key difference was seen with XGBoost: while it achieved the highest cross-validation AUC (0.621), its test AUC fell to 0.599. This drop-off, which was not observed in the LR or RF models (both showing a minimal delta of 0.005), suggests that XGBoost was more sensitive to the training data and experienced mild overfitting despite our class-weighting adjustments.

**Table 2.**
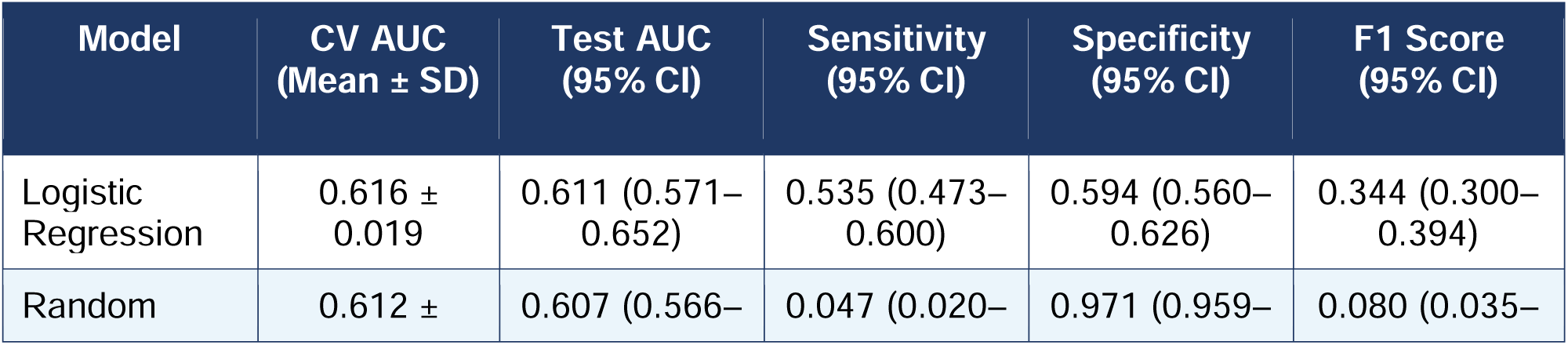
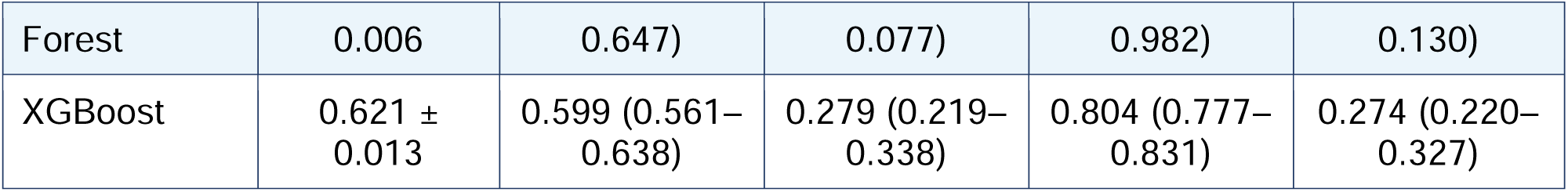
Model performance on the held-out test set with bootstrap 95% confidence intervals (1,000 iterations). CV AUC = mean cross-validation AUC across 5 stratified folds.

| Model | CV AUC<br>(Mean ± SD) | Test AUC<br>(95% CI) | Sensitivity<br>(95% CI) | Specificity<br>(95% CI) | F1 Score<br>(95% CI) |
| --- | --- | --- | --- | --- | --- |
| Logistic Regression | 0.616 ± 0.019 | 0.611 (0.571–0.652) | 0.535 (0.473–0.600) | 0.594 (0.560–0.626) | 0.344 (0.300–0.394) |
| Random | 0.612 ± | 0.607 (0.566– | 0.047 (0.020– | 0.971 (0.959– | 0.080 (0.035– |
| Forest | 0.006 | 0.647) | 0.077) | 0.982) | 0.130) |
| XGBoost | 0.621 ± 0.013 | 0.599 (0.561–0.638) | 0.279 (0.219–0.338) | 0.804 (0.777–0.831) | 0.274 (0.220–0.327) |

### Test Set Performance

On the held-out test set, LR achieved the highest AUC: 0.611 (95% CI: 0.571–0.652), followed by RF: 0.607 (95% CI: 0.566–0.647), and XGBoost: 0.599 (95% CI: 0.561–0.638). No model demonstrated a statistically distinguishable advantage over the others at this sample size. ROC curves for all three classifiers are presented in **Figure 1**; all three curves consistently exceed the no-skill diagonal reference, confirming meaningful discrimination above chance.

**Figure 1.**
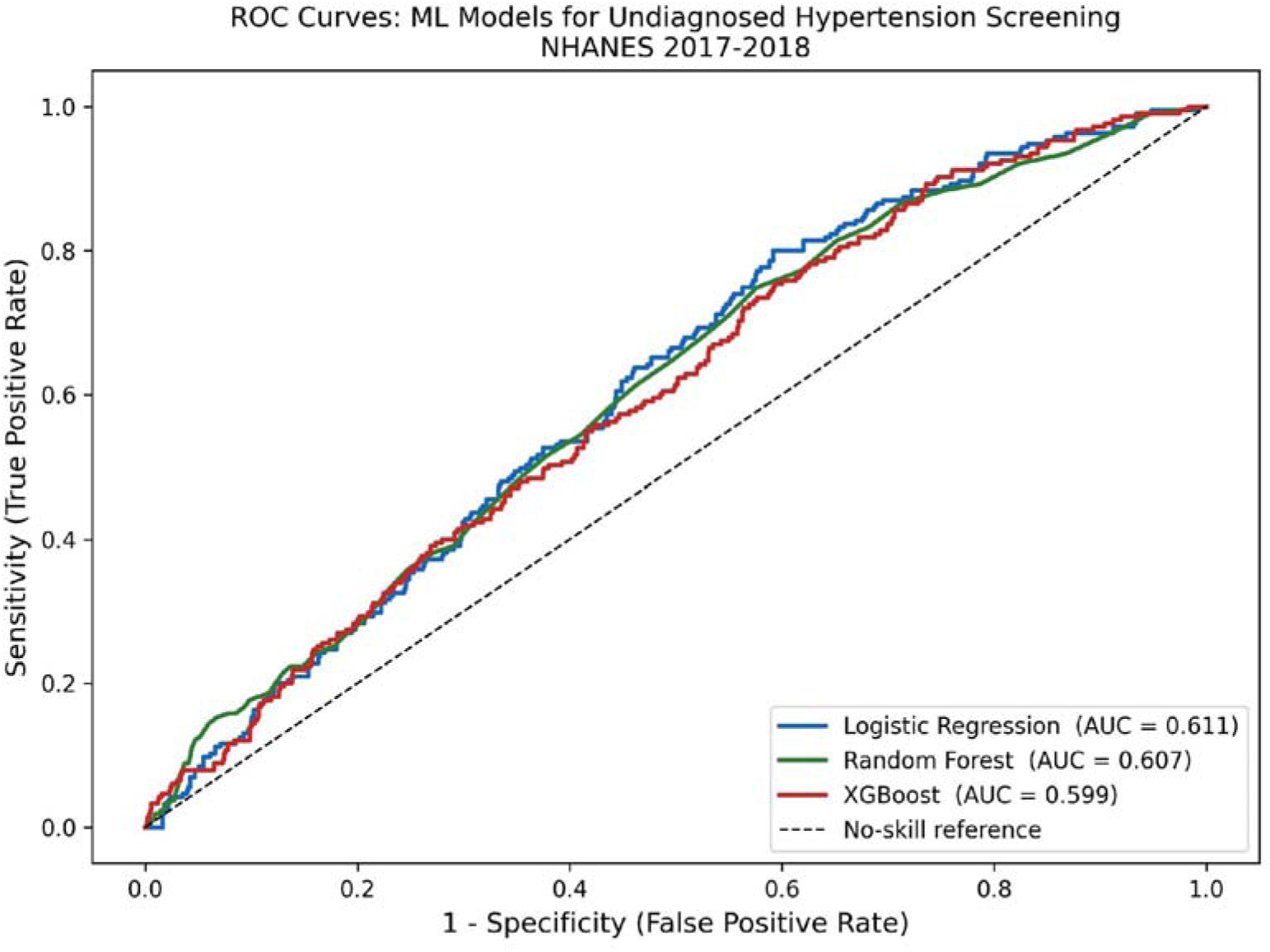
Receiver operating characteristic (ROC) curves for Logistic Regression (AUC=0.611), Random Forest (AUC=0.607), and XGBoost (AUC=0.599) on the held-out test set. Dashed diagonal = no-skill reference line. NHANES 2017–2018.

Substantial differences emerged in sensitivity-specificity profiles. LR demonstrated the most balanced performance: sensitivity 0.535 (95% CI: 0.473–0.600), specificity 0.594 (95% CI: 0.560–0.626), and F1 score 0.344 (95% CI: 0.300–0.394). RF achieved near-perfect specificity (0.971, 95% CI: 0.959–0.982) at the cost of near-absent sensitivity (0.047, 95% CI: 0.020–0.077), rendering it clinically unsuitable as a screening instrument at the default decision threshold. XGBoost occupied an intermediate position, with sensitivity 0.279 (95% CI: 0.219–0.338) and specificity 0.804 (95% CI: 0.777–0.831). Complete performance metrics with 95% CIs are presented in Table 2. These findings highlight a nuanced trade-off relationship between sensitivity and specificity. Tree-based ensembles (RF and XGBoost) favoured specificity at the expense of sensitivity, whereas Logistic Regression maintained a more even balance between the two — the property most relevant to a screening application.

### Feature Importance

Permutation feature importance analysis of the LR model identified **Diabetes Status** as the single most influential predictor (mean AUC decrease ≈ 0.060), followed by **Sex** (≈ 0.035), **Age** (≈ 0.035), and **BMI** (≈ 0.027). Smoking Status, Waist Circumference, Physical Activity, and Sleep Duration each contributed negligibly to model discrimination. Feature importance results are presented in **Figure 2**.

**Figure 2.**
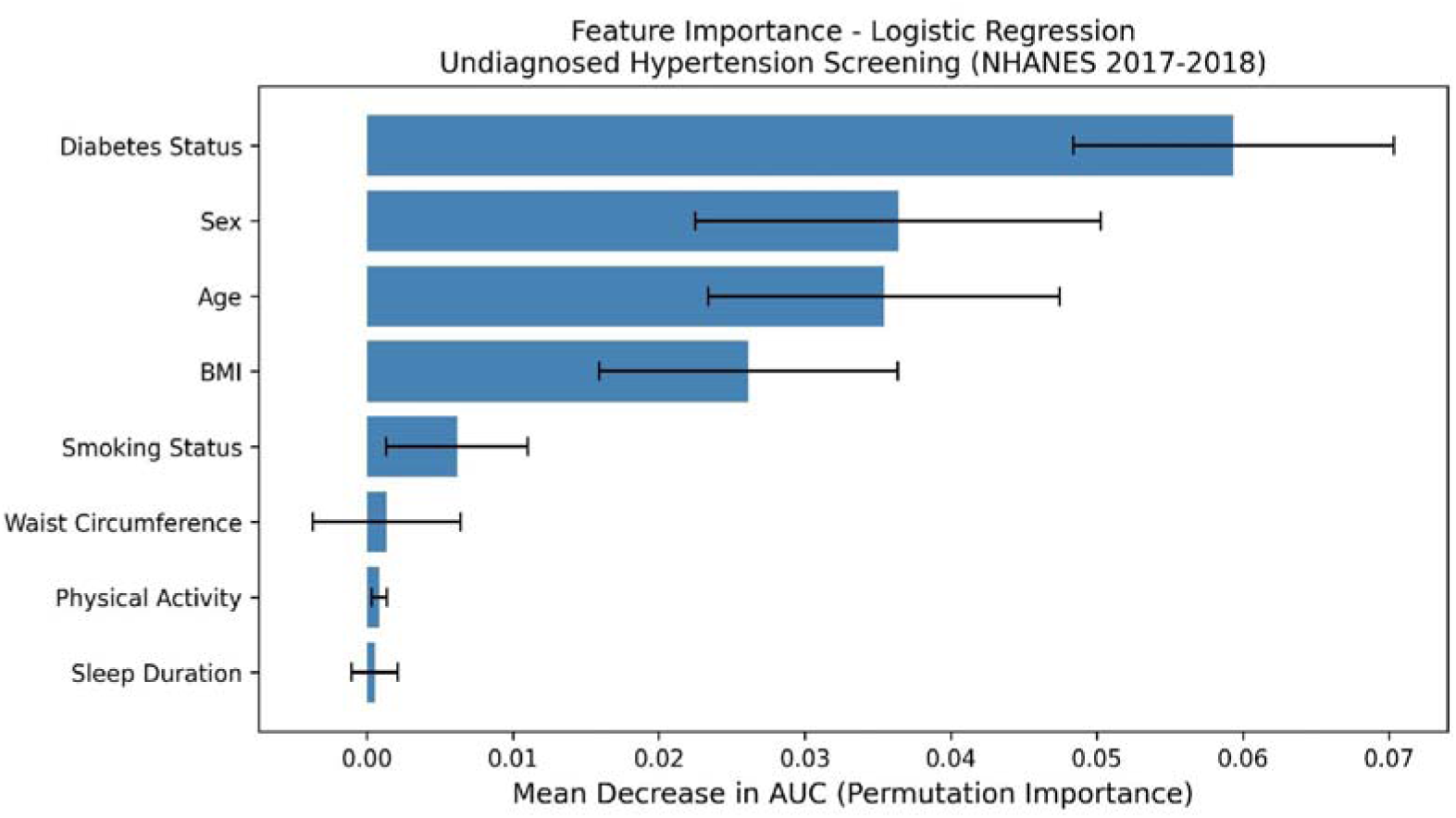
Permutation feature importance for the Logistic Regression model (best-performing by test AUC). Bars represent the mean decrease in AUC when each feature is randomly permuted across 30 independent repeats. Error bars denote ± standard deviation. NHANES 2017–2018.

**Figure 3.**
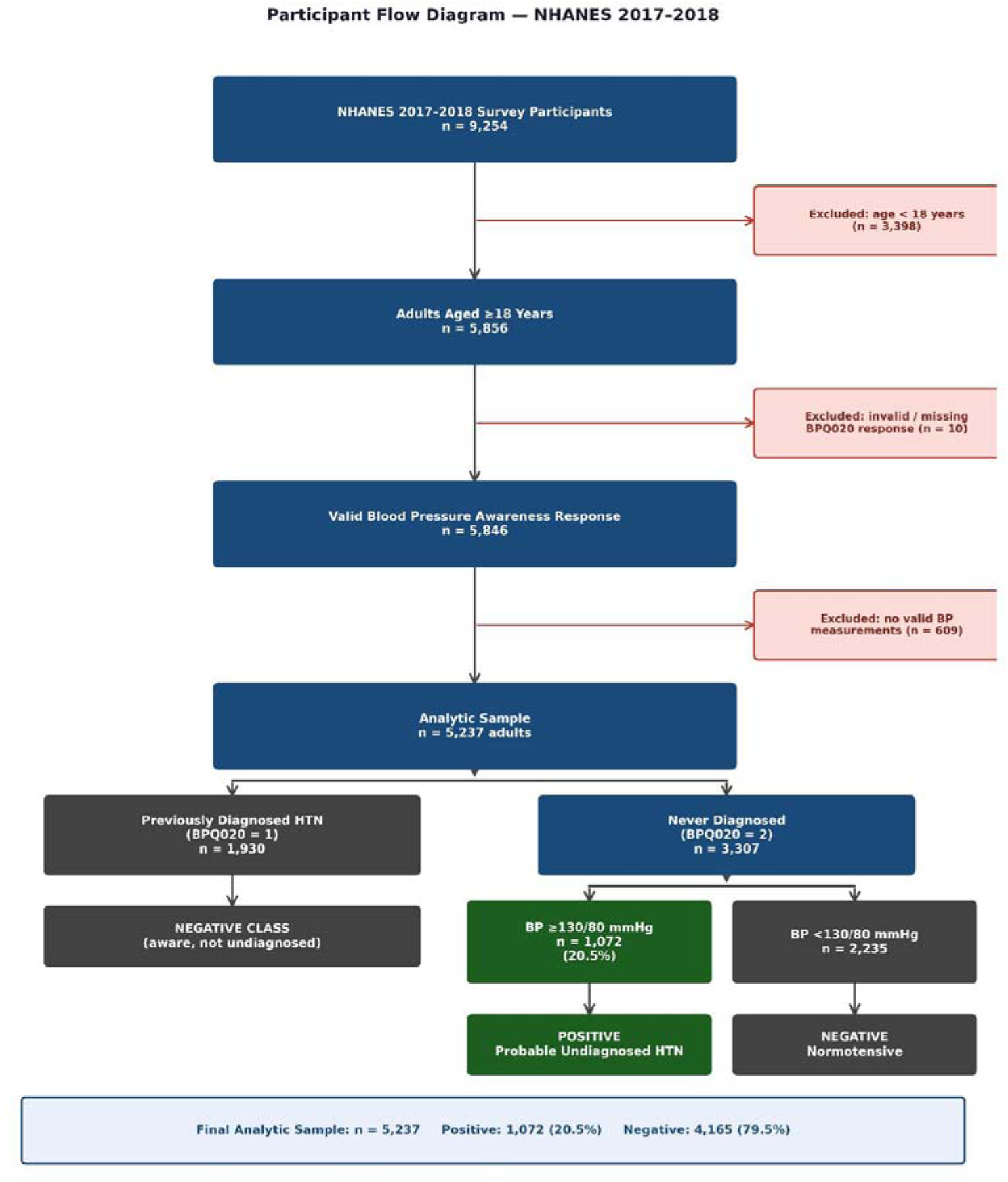
Participant selection flow diagram. Previously diagnosed hypertensives (BPQ020=1; n=1,930) were retained as negative controls as their hypertension status was known and not classifiable as undiagnosed. NHANES = National Health and Nutrition Examination Survey; HTN = hypertension; BP = blood pressure; ACC/AHA = American College of Cardiology/American Heart Association.

## DISCUSSION

### Principal Findings

In this cross-sectional study of 5,237 US adults from NHANES 2017–2018, three machine learning classifiers trained on eight non-invasive variables demonstrated consistent, though modest, discrimination for probable undiagnosed hypertension (AUC range: 0.599–0.611). Logistic Regression (LR) proved to be the most practical model, striking the best balance between correctly identifying true positives (sensitivity 0.535) and correctly ruling out true negatives (specificity 0.594), in addition to the highest test AUC (0.611). Our analysis showed that a person’s diabetes status, sex, and age were the most important indicators for the best-performing model — LR. These findings support the biological and clinical plausibility of the model and suggest that this method could be a straightforward way to screen for hypertension in community health programs.

### Comparison with Existing Literature

Our findings are contextually consistent with prior NHANES-based ML studies on hypertension prediction. López-Martínez et al. (2020) reported an AUC of 0.77 for an artificial neural network predicting hypertension in NHANES using non-invasive features;^4^ however, their model included a broader variable set and targeted prevalent hypertension rather than the specific undiagnosed subset. Guo et al. (2023) applied 13 ML classifiers to NHANES 2017–2020 data, incorporating laboratory variables, and achieved superior discrimination.^5^ The superior discrimination (higher accuracy) achieved by Guo et al. is largely attributable to the inclusion of laboratory variables, whereas our study deliberately restricted the feature set to eight non-invasive predictors to maximize clinical utility in resource-constrained settings. Zhao et al. (2021) reported an AUC of 0.92 for RF using non-invasive features from a Chinese physical examination cohort;^10^ the substantial performance gap likely reflects population-specific characteristics and inclusion of a broader set of predictor variables. Hung et al. (2021) applied ML to masked hypertension prediction — a concept related to but distinct from undiagnosed hypertension — achieving an AUC of 0.837 using LR, RF, XGBoost, and ANN;^11^ our study targets the clinically distinct and arguably more urgent phenotype of completely undiagnosed hypertension. The most comprehensive recent benchmark, Amirian and Zarpoosh (2025), achieved an AUC of 0.68–0.73 in an Iranian cohort with 38 variables, including laboratory investigations, externally validated on NHANES;^12^ our AUC of 0.611 with eight non-invasive variables is commensurate with what can be achieved when laboratory access is deliberately excluded as a design constraint.

### Interpretability and Clinical Meaning

The prominence of Diabetes Status as the most influential predictor is counterintuitive and requires careful explanation. Rather than indicating that diabetics are more likely to have undiagnosed hypertension, Table 1 data reveal the opposite: participants with diabetes had markedly lower rates of probable undiagnosed HTN (10.1%) compared to those without diabetes (16.4%; p<0.001). This finding is explained by differential healthcare contact: individuals with diabetes attend clinical services more frequently, increasing the likelihood that any coexisting hypertension has already been diagnosed and therefore classified as a negative control. The model uses diabetes status as an inverse marker of diagnostic unawareness, reflecting healthcare engagement patterns rather than a direct biological mechanism.

Male predominance in the undiagnosed positive class (56.5% vs. 46.5% in the negative class; p<0.001) is consistent with prior reports of lower healthcare utilisation and less frequent blood pressure monitoring among men.^13^

RF’s near-absent sensitivity (0.047) despite an adequate AUC (0.607) demonstrates a known limitation of tree-based ensembles operating at the default threshold on imbalanced data. At the default 0.5 threshold, the model turns toward the majority class (given the class imbalance), despite the weighting applied during training — a pattern that inflates apparent discrimination (AUC) while suppressing the detection of true positive cases.

XGBoost demonstrated the highest cross-validation AUC (0.621) but the lowest test AUC (0.599). This discrepancy is consistent with mild overfitting, which is not fully remediated by the class weighting that we applied for XGBoost. For screening instruments where the primary objective is maximising case identification, sensitivity is the priority metric, and threshold optimisation below 0.5 may substantially improve RF sensitivity without materially reducing overall discrimination (AUC score). This way, researchers can significantly improve the clinical utility of models like Random Forest, ensuring that fewer undiagnosed cases are missed in community screening programs.

### Clinical Implications

A screening model relying on eight non-invasive, laboratory-free variables could feasibly be deployed by community health workers, pharmacists, or through self-administered digital questionnaires in settings without clinical infrastructure. This is particularly relevant in South Asian contexts, including Nepal, where laboratory facilities may be geographically or economically inaccessible. While the AUC of 0.611 falls below the threshold typically considered “good” discrimination (≥0.70), it meaningfully exceeds chance performance. At the observed prevalence of 20.5%, the LR model yields an estimated positive predictive value (PPV) of approximately 25% and a negative predictive value (NPV) of approximately 83%, indicating that while three in four positive screens are false positives, the model reliably rules out probable undiagnosed hypertension in negative screens. These characteristics position the model as a first-pass screening instrument for identifying individuals who warrant confirmatory blood pressure measurement, not as a diagnostic tool in its own right.

### Strengths

Our study has a number of methodological strengths. First, by defining our outcome as objective blood pressure elevation in patients who are unaware of their condition, we created a screening target that is more clinically practical than traditional risk prediction models. We also intentionally chose to focus on non-invasive variables, prioritizing the ease of applicability of this model in real-world settings rather than chasing lab values for maximum performance metrics. Next, we used the 2017 ACC/AHA threshold to ensure our screening captures a broader at-risk population compared to older guidelines. We have also emphasized scientific transparency and reproducibility by providing a TRIPOD-compliant participant flow diagram, p-values for all baseline comparisons, using bootstrap confidence intervals for all metrics, and making our full analysis pipeline available upon request to the corresponding author.

### Limitations

Several limitations must be acknowledged in our study. The cross-sectional design precludes causal inference and cannot establish the temporal precedence of risk factors before hypertension onset. No external validation in an independent cohort was performed, limiting generalisability claims. NHANES uses complex multistage probability sampling; sampling weights were not applied to the ML pipeline, and population-level prevalence estimates should be interpreted with caution. Key predictor variables — including smoking, physical activity, diabetes status, and sleep duration — relied on self-report, introducing potential recall and social desirability bias. Physical activity was operationalised using vigorous work activity (PAQ605) rather than leisure-time physical activity (PAQ620); this distinction may limit comparability with cardiovascular risk studies that preferentially use recreational activity measures. Formal hyperparameter optimization was not performed; default configurations were used throughout. Calibration curves, decision curve analysis, and SHAP-based directional interpretability were not implemented, representing methodological areas for future refinement.

### Future Directions

External validation in independent cohorts — including South Asian, African, and European population datasets — is the most critical immediate priority. Threshold optimization for tree-based classifiers, SHAP-based directional importance analysis, integration of socioeconomic and dietary variables, and calibration assessment represent enhancements that may meaningfully improve both performance and equity. Prospective validation in community health worker settings would establish the real-world utility of this screening approach.

## CONCLUSION

In this cross-sectional study of 5,237 US adults from NHANES 2017–2018, ML classifiers trained exclusively on eight non-invasive variables achieved modest but consistent discrimination for identifying probable undiagnosed hypertension. Logistic Regression demonstrated the most clinically balanced performance (AUC 0.611, 95% CI: 0.571–0.652; sensitivity 0.535; specificity 0.594; estimated NPV ∼83%). Diabetes status, sex, and age emerged as the most influential predictors, with diabetes status acting paradoxically as an inverse predictor of unawareness, reflecting differential healthcare engagement. These findings support the feasibility of laboratory-free, non-invasive screening tools for undiagnosed hypertension in community settings. External validation and prospective evaluation are required before clinical implementation.

## DECLARATIONS

### Ethics Approval

This study used publicly available, de-identified NHANES data. No additional ethical approval was required.

### Data Availability

NHANES 2017–2018 data are publicly available at: https://wwwn.cdc.gov/nchs/nhanes/continuousnhanes/default.aspx?BeginY ear=2017

### Code Availability

The complete analysis pipeline will be made available on request to the corresponding author.

## Acknowledgements

This project is internally referred to by the lead author as ‘*Project AbhiManyu.’ Abhimanyu* was the son of *Arjuna* and was the valiant warrior prince of the great Hindu epic *Mahabharat,* who entered *Chakravyuh* without the knowledge of returning safely. It represents an initial step toward integrating machine learning methodology into clinical research training for early-career physicians in resource-limited settings.

